# A Novel Triage Tool of Artificial Intelligence-Assisted Diagnosis Aid System for Suspected COVID-19 Pneumonia in Fever Clinics

**DOI:** 10.1101/2020.03.19.20039099

**Authors:** Cong Feng, Lili Wang, Xin Chen, Yongzhi Zhai, Feng Zhu, Hua Chen, Yingchan Wang, Xiangzheng Su, Sai Huang, Lin Tian, Weixiu Zhu, Wenzheng Sun, Liping Zhang, Qingru Han, Juan Zhang, Fei Pan, Li Chen, Zhihong Zhu, Hongju Xiao, Yu Liu, Gang Liu, Wei Chen, Tanshi Li

**Author notes:** Equal contribution to this work as co-first authors (Cong Feng, Lili Wang, Xin Chen). To whom the correspondence should be addressed. **Corresponding Author:** Tanshi Li,. Fever clinic of the Emergency Department, First Medical Center, General Hospital of People’s Liberation Army, Beijing, 100853, China., Wei Chen,. Fever clinic of the Emergency Department, First Medical Center, General Hospital of People’s Liberation Army, Beijing, 100853, China. **Funding:** There is no financial funding or interest to report.

## Abstract

**Background:** Currently, the prevention and control of the novel coronavirus disease (COVID-19) outside Hubei province in China, and other countries have become more and more critically serious. We developed and validated a diagnosis aid model without computed tomography (CT) images for early identification of suspected COVID-19 pneumonia (S-COVID-19-P) on admission in adult fever patients and made the validated model available via an online triage calculator.

**Methods:** Patients admitted from Jan 14 to February 26, 2020 with the epidemiological history of exposure to COVID-19 were included [Model development (n = 132) and validation (n = 32)]. Candidate features included clinical symptoms, routine laboratory tests, and other clinical information on admission. Features selection and model development were based on the least absolute shrinkage and selection operator (LASSO) regression. The primary outcome was the development and validation of a diagnostic aid model for S-COVID-19-P early identification on admission.

**Results:** The development cohort contained 26 S-COVID-19-P and 7 confirmed COVID-19 pneumonia cases. The final selected features included 1 variable of demographic information, 4 variables of vital signs, 5 variables of blood routine values, 7 variables of clinical signs and symptoms, and 1 infection-related biomarker. The model performance in the testing set and the validation cohort resulted in the area under the receiver operating characteristic (ROC) curves (AUCs) of 0.841 and 0.938, the F-1 score of 0.571 and 0.667, the recall of 1.000 and 1.000, the specificity of 0.727 and 0.778, and the precision of 0.400 and 0.500. The top 5 most important features were Age, IL-6, SYS_BP, MONO%, and Fever classification. Based on this model, an optimized strategy for S-COVID-19-P early identification in fever clinics has also been designed.

**Conclusions:** S-COVID-19-P could be identified early by a machine-learning model only used collected clinical information without CT images on admission in fever clinics with a 100% recall score. The well-performed and validated model has been deployed as an online triage tool, which is available at https://intensivecare.shinyapps.io/COVID19/.

## Introduction

Since December 2019, the outbreak of novel coronavirus disease (COVID-19; previously known as 2019-nCoV) (1), which causes severe pneumonia and acute respiratory syndrome (2-5). Until February 29^th^, 2020, the total reported confirmed COVID-19 pneumonia (C-COVID-19-P) cases have reached 85,403, including 79,394 in China and 6,009 in other countries, and the number of cases is increasing rapidly and internationally (6, 7).

The main reason for the outbreak of infected cases in the early stage of the epidemic was the inability to rapidly and effectively detect such a large number of suspected cases(8). Outside Hubei Province, such as Beijing, with a large population, sporadic and clustered cases have continuously been reported. Some other countries and regions, prominently in South Korea, Japan, Iran, etc., are reporting more and more confirmed cases (4, 6, 9, 10). Currently, epidemic prevention and control outside Hubei province and other countries have become more and more critically serious. Therefore, establishing an early identification method of suspected COVID-19 pneumonia (S-COVID-19-P) and optimizing triage strategies for fever clinics is urgent and essential for the coming global challenge.

The identification of S-COVID-19-P relies on the following criteria: the epidemiological history, clinical signs and symptoms, routine laboratory tests (such as lymphopenia), and positive Chest computerized tomography (CT) findings(3). However, clinical symptoms and routine laboratory tests are sometimes non-specific(2, 3). Although CT is becoming a major diagnostic tool help for early screening of S-COVID-19-P, the resources of the designated CT room were relatively limited, especially in less-developed regions and when the influx of patients substantially outweighed the medical service capacities in fever clinics(11, 12). Moreover, not all patients with clinical symptoms or abnormal blood routine values need CT examination, except radiation injury, high cost, and other restrictions. Therefore, it is critical to integrate and take the advantages of clinical signs and symptoms, routine laboratory tests, and other clinical information on admission before further CT examination, which would strengthen the ability of early identification of S-COVID-19-P, improve the triage strategies in fever clinics and make a balance between standard medical principles and limited medical resources.

The increase of secondary analysis in the emergency departments and intensive care units has given feasibility to get ‘real-time’ data from the electronic medical records, thus making them enable for ‘real world’ research (13, 14). This term pertains to machine-learning algorithms to analyze specific clinical cohorts and develop models for diagnosis aid or decision support in emergent triage(15). Such models could be a cost-effective assisting tool to integrate clinical signs and symptoms, blood routine values, and infection-related biomarkers on admission for S-COVID-19-P early identification(16-18).

The aim of this study was the development and validation of a diagnostic aid model on admission without CT images for early identification of S-COVID-19-P in adult fever patients with the epidemiological history of exposure to COVID-19. The model performance was also compared to some infection-related biomarkers on admission in the general population admitted to the fever clinic. The well-performed model is available as an online triage calculator, and based on it, the optimized strategy for S-COVID-19-P early identification in fever clinics has also been discussed. We present the following article in accordance with the STROBE reporting checklist

## Methods

### Ethical Statement

The study was conducted in accordance with the Declaration of Helsinki (as revised in 2013). The study was approved by institutional ethics committee of the General Hospital of the PLA (NO.: 2020-094 the registration number of ethics board). This study was based on the retrospective and secondary analysis of the clinical data. Medical records collection was passive and had no impact on patient safety. Studies performed on de-identified data constitute non-human subject research, thus no informed consent was required for this study. The authors are accountable for all aspects of the work in ensuring that questions related to the accuracy or integrity of any part of the work are appropriately investigated and resolved.

### Study design and population: development and validation cohorts

We developed a novel diagnosis aid model for early identification S-COVID-19-P based on the retrospective analysis of a single-center study. All patients admitted to the fever clinic of the emergency department of the First medical center, Chinese People’s Liberation Army General Hospital (PLAGH) in Beijing with the epidemiological history of exposure to COVID-19 according to WHO interim guidance was enrolled in this study. The fever clinic is a department for adults (*i*.*e*., aged ≥14 years) specializing in the identification of infectious diseases, especially for S-COVID-19-P. We recruited patients from Jan 14 to Feb 9, 2020, as a model development cohort. Meanwhile, we also recruited patients from Feb 10 to Feb 26, 2020, as a dataset for model validation.

The study was conducted in accordance with the Declaration of Helsinki (as revised in 2013). Data collection was passive and had no impact on patient safety. Studies performed on de-identified data constitute non-human subject research, thus no institutional or ethical approvals were required for this study.

### The definition of S-COVID-19-P

All recruited patients on admission were given vital signs, blood routine, infection-related biomarkers, influenza viruses (A+B), and chest CT examination. The patients who have the epidemiological history and CT imaging characteristics of viral pneumonia and any other one of the following two clinical signs were diagnosed as S-COVID-19-P, which according to the “Guidelines for diagnosis and management of novel coronavirus pneumonia (The sixth Edition)” published by Chinese National Health and Health Commission on Feb 18, 2020 (6th-Guidelines-CNHHC). The two clinical signs including 1) Fever and/or respiratory symptoms; 2) Total count of leukocyte was normal or decreased, or lymphopenia (<1.0× 10_9_/L).

### The definition of C-COVID-19-P

Throat swab specimens from the upper respiratory tract were obtained from all patients on admission and then maintained in the viral-transport medium. Those with positive results were clinically identified as C-COVID-19-P(3). Laboratory confirmation of COVID-19 infection was done in four different institutions: the PLAGH, the Haidian District Disease Control and Prevention (CDC) of Beijing, the Beijing CDC, and the Academy of Military Medical Sciences. COVID-19 infection was confirmed by real-time RT-PCR using the same protocol described previously(2). RT-PCR detection reagents were provided by the four institutions.

### Data extraction

All data of each patient were extracted on admission, which included demographic information, comorbidities, epidemiological history of exposure to COVID-19, vital sign, blood routine values, clinical symptoms, infection-related biomarkers, influenza viruses (A+B) test, CT findings, and days of illness onset to the first admission. All data were checked and missing data were obtained through direct communication with the other two attending doctors (XC and YZ).

### Outcomes

The primary outcome is the development and validation of a diagnostic aid model for S-COVID-19-P early identification on admission. The secondary outcome is the comparison of the diagnostic performance between the diagnosis aid model and infection-related biomarkers on admission.

### Diagnosis aid model and candidate features

For early identification of S-COVID-19-P on admission, a diagnostic aid model that only used clinical information and based on the availability of patients’ medical records was developed. We included following candidate features: 1) 2 variables of demographic information (*e*.*g*., age and gender); 2) 4 variables of vital signs (*e*.*g*., temperature, heart rate, etc.); 3) 20 variables with blood routine values (*e*.*g*., white blood cell count, red blood cell count, hemoglobin, hematocrit, etc.); 4) 17 variables of clinical signs and symptoms [e.g., fever, fever classification (°C, normal: <= 37.0, mild fever: 37.1-38.0, moderate fever: 38.1-39.0, severe fever: >=39.1), cough, muscle ache, etc.]; 5) 2 infection-related biomarkers (*e*.*g*., C-reactive protein and Interleukin-6); 6) 1 other variable: days from illness onset to first admission (DOA). The complete candidate features list is shown in Table 1.

**Table 1:**
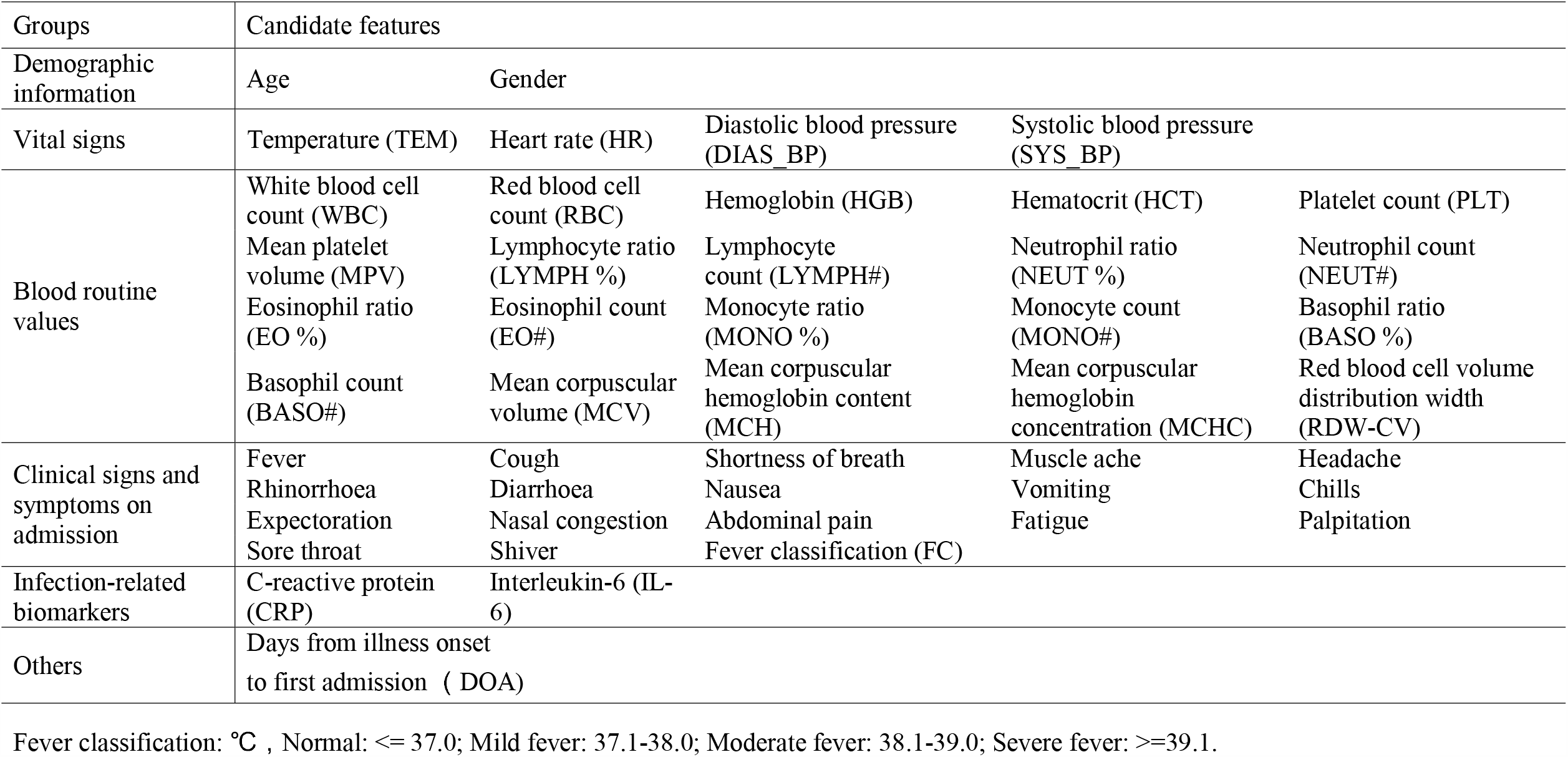
Candidate features for diagnosis aid model

| Groups | Candidate features |  |  |  |  |
| --- | --- | --- | --- | --- | --- |
| Demographic information | Age | Gender |  |  |  |
| Vital signs | Temperature (TEM) | Heart rate (HR) | Diastolic blood pressure (DIAS_BP) | Systolic blood pressure (SYS_BP) |  |
| Blood routine values | White blood cell count (WBC) | Red blood cell count (RBC) | Hemoglobin (HGB) | Hematocrit (HCT) | Platelet count (PLT) |
|  | Mean platelet volume (MPV) | Lymphocyte ratio (LYMPH %) | Lymphocyte count (LYMPH#) | Neutrophil ratio (NEUT %) | Neutrophil count (NEUT#) |
|  | Eosinophil ratio (EO %) | Eosinophil count (EO#) | Monocyte ratio (MONO %) | Monocyte count (MONO#) | Basophil ratio (BASO %) |
|  | Basophil count (BASO#) | Mean corpuscular volume (MCV) | Mean corpuscular hemoglobin content (MCH) | Mean corpuscular hemoglobin concentration (MCHC) | Red blood cell volume distribution width (RDW-CV) |
| Clinical signs and symptoms on admission | Fever<br>Rhinorrhoea<br>Expectoration<br>Sore throat | Cough<br>Diarrhoea<br>Nasal congestion<br>Shiver | Shortness of breath<br>Nausea<br>Abdominal pain<br>Fever classification (FC) | Muscle ache<br>Vomiting<br>Fatigue | Headache<br>Chills<br>Palpitation |
| Infection-related biomarkers | C-reactive protein (CRP) | Interleukin-6 (IL-6) |  |  |  |
| Others | Days from illness onset to first admission ( DOA) |  |  |  |  |

Fever classification: °C , Normal: <= 37.0; Mild fever: 37.1-38.0; Moderate fever: 38.1-39.0; Severe fever: >=39.1.

### Features selection and model development

Candidate features were selected based on expert opinion and availability of the medical records. For the model, we compared 4 different algorithms: 1) logistic regression with the least absolute shrinkage and selection operator (LASSO), 2) logistic regression with Ridge regularization, 3) decision tree, 4) Adaboost algorithms, and found that logistic regression with LASSO achieved overall best performances in both the testing set and external validation set in terms of AUC and recall score (Table S1). Features selection and model development were performed in the development cohort only and using logistic regression with LASSO regularization (LASSO regression) which is one of the models that shrinks some regression coefficients toward zero, thereby effectively select important features and improve the interpretability of the model(19). The feature selection and model development were performed in Python 3.7. During the model training, we randomly held out 20% of the cohort data as a testing set and then used 10-fold cross-validation to yield the optimal of the LASSO regularization parameter in the training and validation sets. All features were normalized to standard uniform distribution according to the training and validation sets and then this transformation was applied to both the held-out testing set as well as the external validation set. All computations were achieved by scikit-learn (version: 0.22.1) in python. Random oversampling was performed to construct balanced data on training and validation sets by using the imblearn python package (version 0.6.2).

### Model validation

After model development, we used the cohort with the epidemiological history from February 10 to February 26, 2020, for model validation. The model validation was also performed in python.

### Features Importance Ranking

Feature importance was performed in the development cohort. The associated coefficient weights corresponding to the logistic regression model were used to identify and rank the feature importance.

### Comparison of diagnostic performance among diagnosis aid model and infection-related biomarkers

Lymphocyte count (LYMPH#), C-reactive protein (CRP), and Interleukin-6 (IL-6) were evaluated on admission. Lymphopenia (<1.0×10^9^/L) was one of the three diagnostic criteria for S-COVID-19-P according to the 6th-Guidelines-CNHHC. Elevated CRP (>0.8 mg/L) and elevated IL-6 (>5.9 pg/ml) were both important infection-related biomarkers. The diagnostic performance among the diagnosis aid model and biomarkers for early identifying S-COVID-19-P was also compared.

The entire workflow is shown in Figure 1.

**Figure 1.**
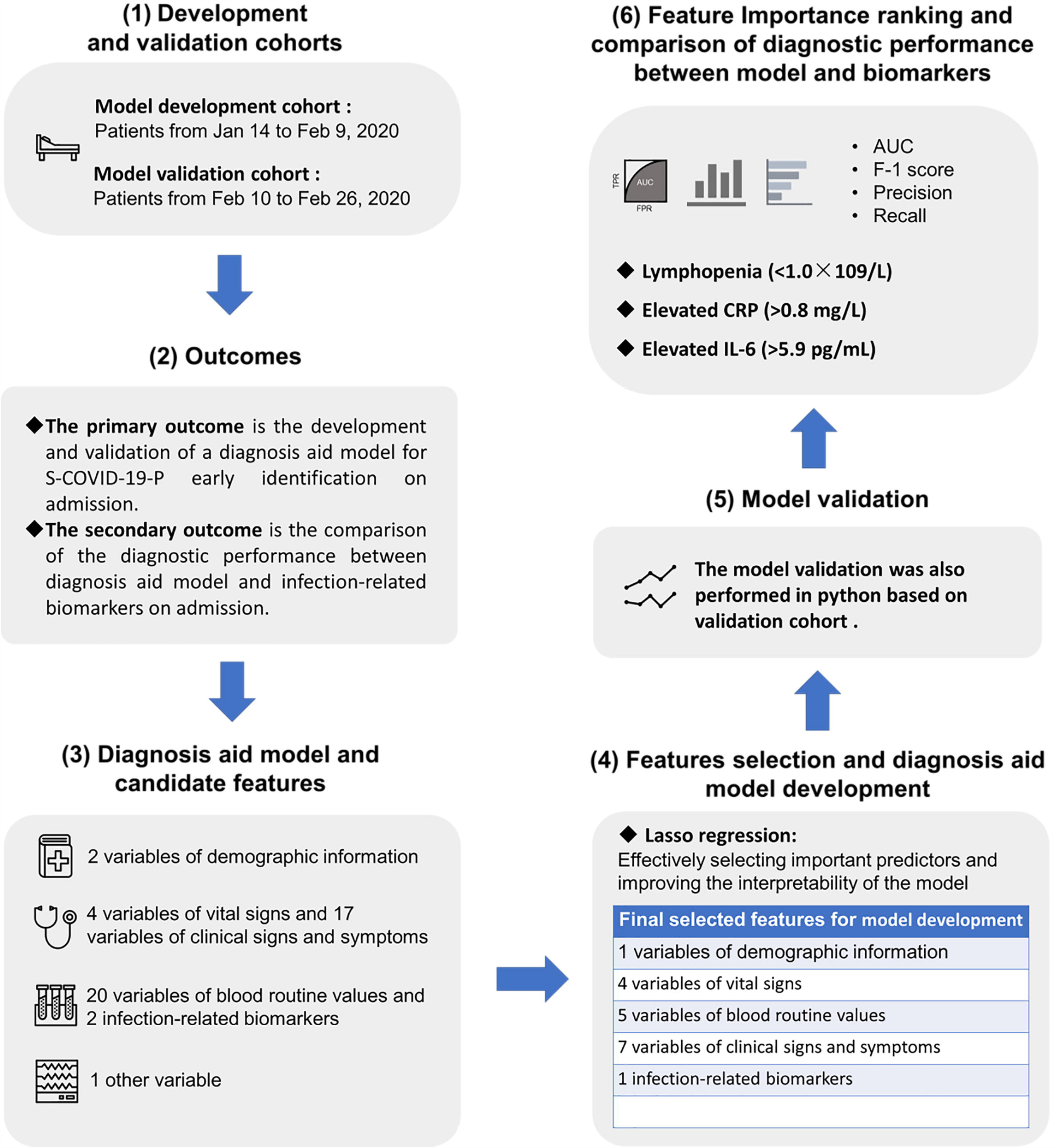
The study overview of the Artificial Intelligence-Assisted Diagnosis Aid System for Suspected COVID-19 Pneumonia, including (1) Development and validation cohorts, (2) Outcomes, (3) Diagnosis aid model and candidate features, (4) Features selection and diagnosis aid model development, (5) Model validation, and (6) Feature Importance ranking and comparison of diagnostic performance between model and biomarker. S-COVID-19-P= suspected COVID-19 pneumonia,

### Statistical Analysis and Performance Evaluation

Continuous variables were expressed as median with interquartile range (IQR) and compared with the Mann-Whitney U test; categorical variables were expressed as absolute (n) and relative (%) frequency and compared by χ^2^ test or Fisher’s exact test. A two-sided α of less than 0.05 was considered statistically significant. Statistical analysis was performed by R version 3.5.1.

Model performance was evaluated by 1) the area under the receiver operating characteristic (ROC) curve (AUC) (20), 2) F-1 score, 3) Precision, 4) Sensitivity (Recall), 5) Specificity. AUC, ranging from 0 to 1, the higher the better, indicates the algorithm’s performances. Precision is the fraction of true positive classification among the positive results classified by the algorithm; higher accuracy indicates that the result of the algorithm is reliable. The Recall is the fraction of true positive classification among all the true samples, which describes the ability to identify true samples (S-COVID-19-P) among the whole population. F1 score is the harmonic average of precision and recall, and a higher F1 score indicates better performance. In this study, to avoid missed suspected cases, recall is the most important reference(21). We considered the model with AUC above 0.80 and recall above 0.95 as the adequate and well-performed model.

## Results

### Study population: development and validation cohorts

In the development cohort, a total of 132 unique admissions with the epidemiological history of exposure to COVID-19 were included from Jan 14 to Feb 9, 2020. 26 patients were clinically identified as S-COVID-19-P according to the 6th-Guidelines-CNHHC and 7 patients out of them were further identified as C-COVID-19-P in Beijing. 10 (38.5%) out of 26 S-COVID-19-P cases were transferred to the CDC after the first laboratory confirmation of COVID-19 infection by PLAGH. The left 16 (61.5%) S-COVID-19-P cases were kept hospitalizing for quarantine and further laboratory confirmation of COVID-19 infection. The 7 C-COVID-19-P cases all belonged to moderate type based on the 6th-Guidelines-CNHHC, no ICU admission, no death occurred, and no excluded patients. (Table 2)

**Table 2:** Demographics, baseline and clinical characteristics of 132 patients admitted to PLA General Hospital (Jan 14–Feb 9, 2020) with the epidemiological history of exposure to COVID-19 in development cohort.

|  | All patients | Non-suspected<br>COVID-19<br>pneumonia<br>cases | Suspected<br>COVID-19<br>pneumonia<br>cases | p-value<sup>1</sup> | Non- confirmed<br>COVID-19<br>pneumonia in<br>suspected cases | Confirmed<br>COVID-19<br>pneumonia in<br>suspected cases | p-value<sup>2</sup> |
| --- | --- | --- | --- | --- | --- | --- | --- |
| Cohort (n) | 132 | 106 | 26 | - | 19 | 7 | - |
| Age, years (median,(IQR)) | 34.0(29.0-42.0) | 33.0(28.0-40.0) | 39.5(36.3-52.3) | 0.004 | 40.0(32.5-54.5) | 39.0(37.0-41.5) | 0.954 |
| Gender (n (%)) |  |  |  | 0.396 |  |  | - |
| Male | 74(56.1%) | 57(53.8%) | 17(65.4%) | - | 12(63.2%) | 5(71.4%) | - |
| Female | 58(43.9%) | 49(46.2%) | 9(34.6%) | - | 7(36.8%) | 2(28.6%) | - |
| Days from illness onset<br>to first admission,<br>(median,(IQR)) | 2.0(1.0-5.0) | 2.0(1.0-5.0) | 2.5(1.0-4.8) | 0.974 | 1.0(1-3.5) | 5.0(3.5-5.5) | 0.017 |
| Comorbidities (n (%)) |  |  |  |  |  |  |  |
| Hypertension | 2(1.5%) | 2(1.9%) | 0(0%) | - | 0(0%) | 0(0%) | - |
| Diabetes | 2(1.5%) | 1(0.9%) | 1(3.8%) | - | 1(5.3%) | 0(0%) | - |
| Cardiovascular disease | 0(0%) | 0(0%) | 0(0%) | - | 0(0%) | 0(0%) | - |
| Chronic obstructive<br>pulmonary disease | 3(2.3%) | 1(0.9%) | 2(7.7%) | - | 2(10.5%) | 0(0%) | - |
| Malignancy | 0(0%) | 0(0%) | 0(0%) | - | 0(0%) | 0(0%) | - |
| Chronic kidney disease | 1(0.8%) | 1(0.9%) | 0(0%) | - | 0(0%) | 0(0%) | - |
| Chronic liver disease | 1(0.8%) | 1(0.9%) | 0(0%) | - | 0(0%) | 0(0%) | - |
| The epidemiological<br>history of exposure to<br>COVID-19 (n (%)) |  |  |  |  |  |  |  |
| History of sojourn or<br>residence (HSR) | 56(42.4%) | 48(45.3%) | 8(30.8%) | 0.263 | 4(21.1%) | 4(57.1%) | 0.149 |
| History of contactation with<br>confirmed COVID-19<br>infected patients (HCCI) | 10(7.6%) | 7(6.6%) | 3(11.5%) | 0.412 | 1(5.3%) | 2(28.6%) | 0.167 |
| History of contactation with persons who had fever or respiratory symptoms (HCFR) | 63(47.7%) | 51(48.1%) | 12(46.2%) | - | 11(57.9%) | 1(14.3%) | 0.081 |
| Clustering onset | 3(2.3%) | 0(0%) | 3(11.5%) | 0.007 | 3(15.8%) | 0(0%) | 0.54 |
| Vital sign on admission |  |  |  |  |  |  |  |
| Heart rate, n/min (median,(IQR)) | 101.5(92.0-112.2) | 99.5(89.5-110.0) | 107.5(100.0-116.2) | 0.035 | 103.0(97.0-122.0) | 110.0(102.5-113.0) | 0.885 |
| Diastolic blood pressure, mmHg (median,(IQR)) | 83.5(75.8-91.0) | 81.0(75.0-88.0) | 89.5(80.5-96.3) | 0.014 | 91.0(79.5-97.0) | 85.0(82.5-90.0) | 0.817 |
| Systolic blood pressure, mmHg (median,(IQR)) | 136.0(125.8-147.2) | 134.0(124.0-143.0) | 145.5(136.2-156.8) | <0.001 | 147.0(138.0-157.5) | 137.0(133.5-152.0) | 0.37 |
| Fever (n (%)) | 93(70.5%) | 70(66.0%) | 23(88.5%) | 0.045 | 17(89.5%) | 6(85.7%) | - |
| Highest temperature, °C (median,(IQR)) | 37.4(36.8-38.0) | 37.4(36.8-37.8) | 37.9(37.4-38.5) | 0.006 | 37.8(37.5-38.3) | 38.5(37.3-38.6) | 0.84 |
| <37.1 | 39(29.5%) | 36(34.0%) | 3(11.5%) | 0.03 | 2(10.5%) | 1(14.3%) | - |
| 37.1–38.0 | 61(46.2%) | 49(46.2%) | 12(46.2%) | - | 10(52.6%) | 2(28.6%) | 0.391 |
| 38.1–39.0 | 27(20.5%) | 18(17.0%) | 9(34.6%) | 0.084 | 5(26.3%) | 4(57.1%) | 0.188 |
| >39.0 | 5(3.8%) | 3(2.8%) | 2(7.7%) | 0.255 | 2(10.5%) | 0(0%) | - |
| Other symptoms on admission (n (%)) |  |  |  |  |  |  |  |
| Cough | 65(59.2%) | 53(50.0%) | 12(46.2%) | 0.895 | 7(36.8%) | 5(71.4%) | 0.19 |
| Shortness of breath | 18(13.6%) | 17(16.0%) | 1(3.8%) | 0.197 | 1(5.3%) | 0(0%) | - |
| Muscle ache | 43(32.6%) | 32(30.2%) | 11(42.3%) | 0.343 | 5(26.3%) | 6(85.7%) | 0.021 |
| Headache | 28(21.2%) | 20(18.9%) | 8(30.8%) | 0.19 | 3(15.8%) | 5(71.4%) | 0.014 |
| Sore throat | 58(43.9%) | 43(40.6%) | 15(57.7%) | 0.175 | 10(52.6%) | 5(71.4%) | 0.658 |
| Rhinorrhoea | 28(21.2%) | 20(18.9%) | 8(30.8%) | 0.19 | 7(36.8%) | 1(14.3%) | 0.375 |
| Diarrhoea | 12(9.1%) | 11(10.4%) | 1(3.8%) | 0.459 | 1(5.3%) | 0(0%) | - |
| Nausea | 4(3.0%) | 3(2.8%) | 1(3.8%) | - | 1(5.3%) | 0(0%) | - |
| Vomiting | 3(2.3%) | 3(2.8%) | 0(0%) | - | 0(0%) | 0(0%) | - |
| Chills | 37(28.0%) | 31(29.2%) | 6(23.1%) | 0.701 | 4(21.1%) | 2(28.6%) | - |
| Shiver | 18(13.6%) | 16(15.1%) | 2(7.7%) | 0.524 | 1(5.3%) | 1(14.3%) | 0.474 |
| Expectoration | 39(29.5%) | 33(31.1%) | 6(23.1%) | 0.481 | 3(15.8%) | 3(42.9%) | 0.293 |
| Abdominal pain | 5(3.8%) | 4(3.8%) | 1(3.8%) | - | 1(5.3%) | 0(0%) | - |
| Fatigue | 44(33.3%) | 37(34.9%) | 7(26.9%) | 0.588 | 4(21.1%) | 3(42.9%) | 0.34 |
| Palpitation | 3(2.3%) | 3(2.8%) | 0(0%) | - | 0(0%) | 0(0%) | - |
| Clinical outcome (n (%)) |  |  |  |  |  |  |  |
| Discharged for home quarantine | 106(80.3%) | 106(100%) | 0(0%) | - | 0(0%) | 0(0%) | - |
| Hospitalization for quarantine | 16(12.1%) | 0(0%) | 16(61.5%) | - | 16(84.2%) | 0(0%) | - |
| Transferred to Disease Control and Prevention (CDC) | 10(7.5%) | 0(0%) | 10(38.5%) | - | 3(15.8%) | 7(100%) | - |
| Death | 0(0%) | 0(0%) | 0(0%) | - | 0(0%) | 0(0%) | - |

Continuous variables were expressed as median with interquartile range (IQR) and compared with the Mann-Whitney U test; categorical variables were expressed as absolute (n) and relative (%) frequency and compared by $\chi^2$ test or Fisher's exact test. A two-sided $\alpha$ of less than 0.05 were considered statistically significant.
COVID-19: 2019 novel coronavirus.
History of sojourn or residence: Within 14 days before the onset of the disease, there was a history of sojourn or residence in the surrounding areas of Wuhan or other confirmed COVID-19 infected case reporting communities.
History of contact with confirmed COVID-19 infected patients: Within 14 days before the onset of the disease, there was a history of contact with confirmed COVID-19 infected patients.
History of contact with persons who had fever or respiratory symptoms: Within 14 days before the onset of the disease, there was a contact history with persons who had fever or respiratory symptoms. The persons come from Wuhan city and its surrounding areas, or come from the community where have reported confirmed COVID-19 infected cases.
P-value1: Suspected COVID-19 pneumonia cases compared to Non-suspected COVID-19 pneumonia cases.
P-value2: Confirmed COVID-19 pneumonia cases compared to Non-confirmed COVID-19 pneumonia in suspected cases.

These S-COVID-19-P cases with a median age of 39.5 (36.3-52.3), 17 (65.4%) were male and the median days of DOA were 2.5 (1.0-4.8). Non-suspected COVID-19 pneumonia (N-S-COVID-19-P) cases with a median age of 33.0 (28.0-40.0), 57 (53.8%) were male and the median days of DOA were 2.0 (1.0-5.0). C-COVID-19-P cases with a median age of 39.0 (37.0-41.5), 5 (71.4%) were male and the median days of DOA were 5.0 (3.5-5.5). (Table 2)

Within 14 days before the onset of the disease, there were 3 (11.5%), 7 (6.6%) and 2 (28.6%) patients had a history of contact with COVID-19 infected patients (laboratory-confirmed infection) in suspected, non-suspected, and confirmed COVID-19 pneumonia cases, respectively. On admission, the median heart rate [107.5 (100.0-116.2) vs 99.5 (89.5-110.0), p=0.035],diastolic blood pressure [89.5 (80.5-96.3) vs 81.0 (75.0-88.0), p=0.014], systolic blood pressure [145.5 (136.2-156.8) vs 134.0 (124.0-143.0), p<0.001] and the highest temperature [37.9 (37.4-38.5) vs 37.4 (36.8-37.8), p=0.006] were much higher in S-COVID-19-P cases than in N-S-COVID-19-P cases. (Table 2)

The most common symptoms at onset of illness were fever [23 (88.5%), 70 (66.0%)], sore throat [15 (57.7%), 43 (40.6%)], and cough [12 (46.2%), 53 (50.0%)) in S-COVID-19-P and N-S-COVID-19-P cases, respectively. However, in C-COVID-19-P cases, muscle ache 6 (85.7%) and headache 5 (71.4%) were also the most common symptoms besides the fever 6 (85.7%), cough 5 (71.4%), and sore throat 5 (71.4%). (Table 2)

The blood routine values of patients on admission showed lymphopenia [lymphocyte count<1·0 × 10^9^/L; 9 (34.6%), 17 (16.0%) and 1 (14.3%)] and elevated monocyte ratio [monocyte ratio > 0.08; 12 (46.2%), 18 (17.0%) and 4 (57.1%)] in S-COVID-19-P, N-S-COVID-19-P and C-COVID-19-P cases, respectively. Early lymphopenia (p=0.051) and the elevated monocyte ratio (p=0.003) were more prominent in S-COVID-19-P than N-S-COVID-19-P cases, but no statistically difference between C-COVID-19-P and non-C-COVID-19-P in S-COVID-19-P cases. The ratio of elevated CRP cases on admission was more in S-COVID-19-P cases than N-S-COVID-19-P cases [13(50.0%) vs 29(27.4%), p=0.035], but no statistical significance between C-COVID-19-P cases and non-C-COVID-19-P in S-COVID-19-P cases [6(85.7%) vs 7(36.8%),p=0.190]. The ratio of elevated IL-6 cases on admission was also more in S-COVID-19-P cases than N-S-COVID-19-P cases [16(61.5%) vs 34(32.1%), p=0.007], but no statistical significance between C-COVID-19-P cases and non-C-COVID-19-P in S-COVID-19-P cases [6(85.7%) vs 10(52.6%), p=0.190]. (Table 3)

**Table 3:**
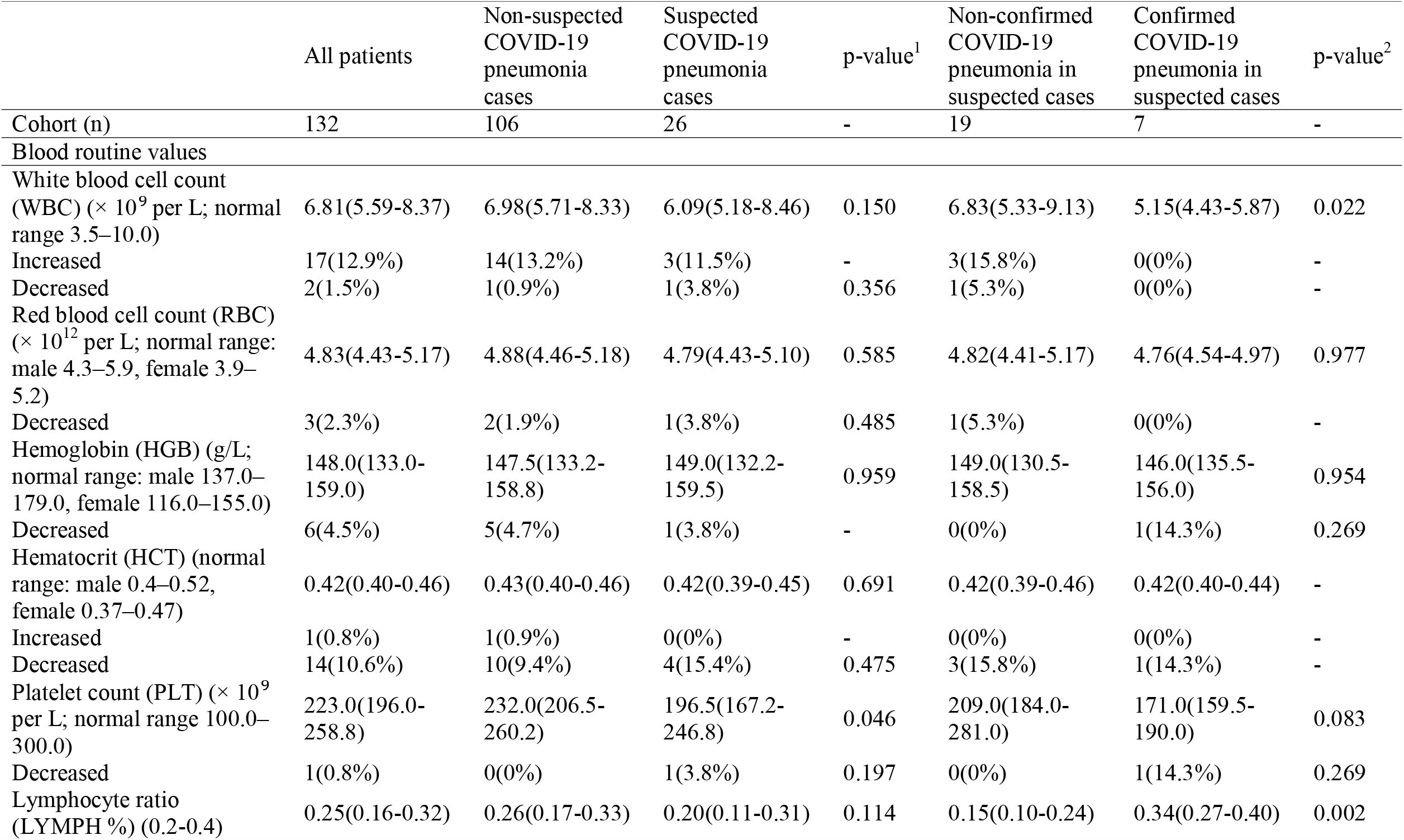
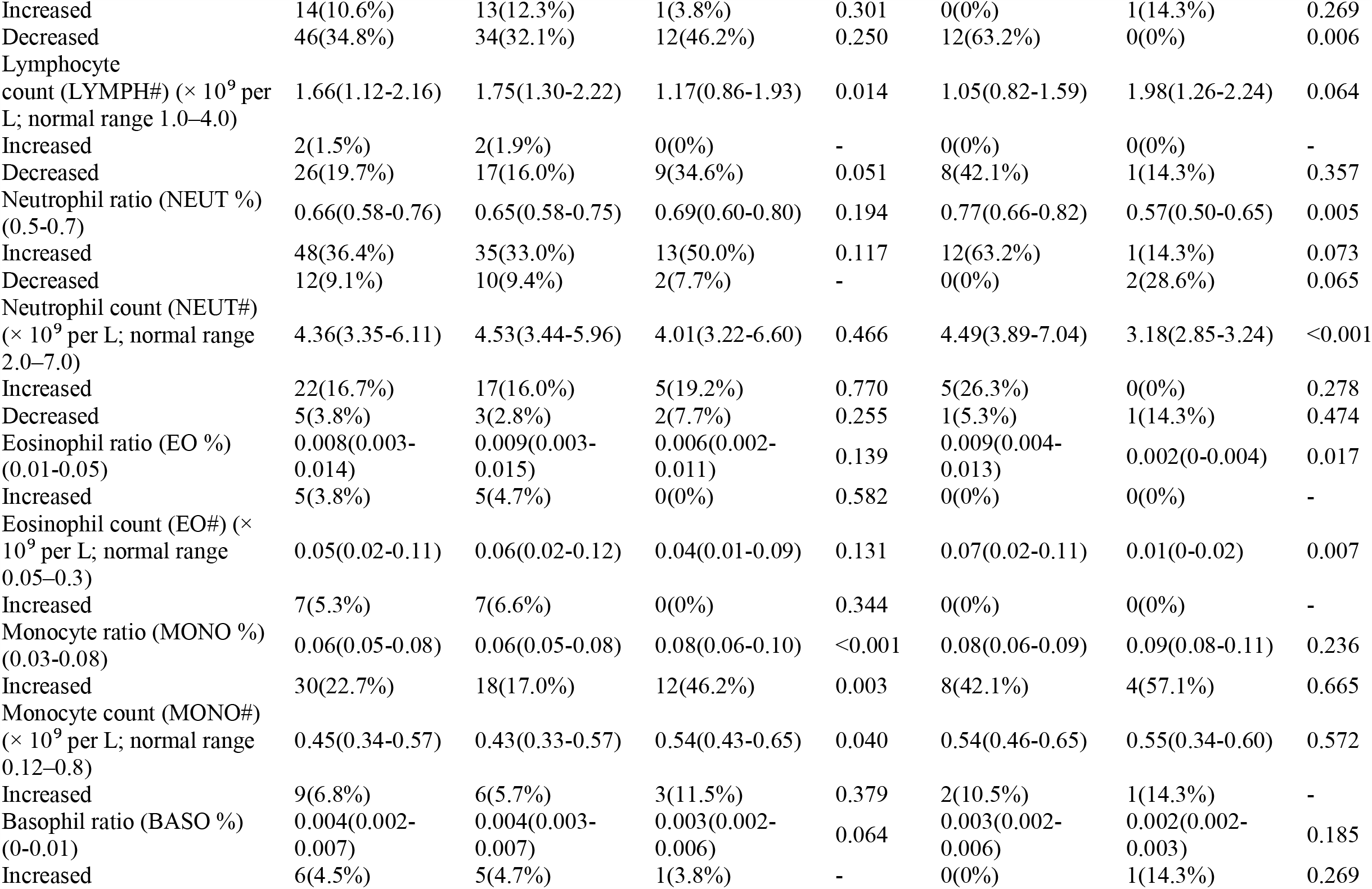
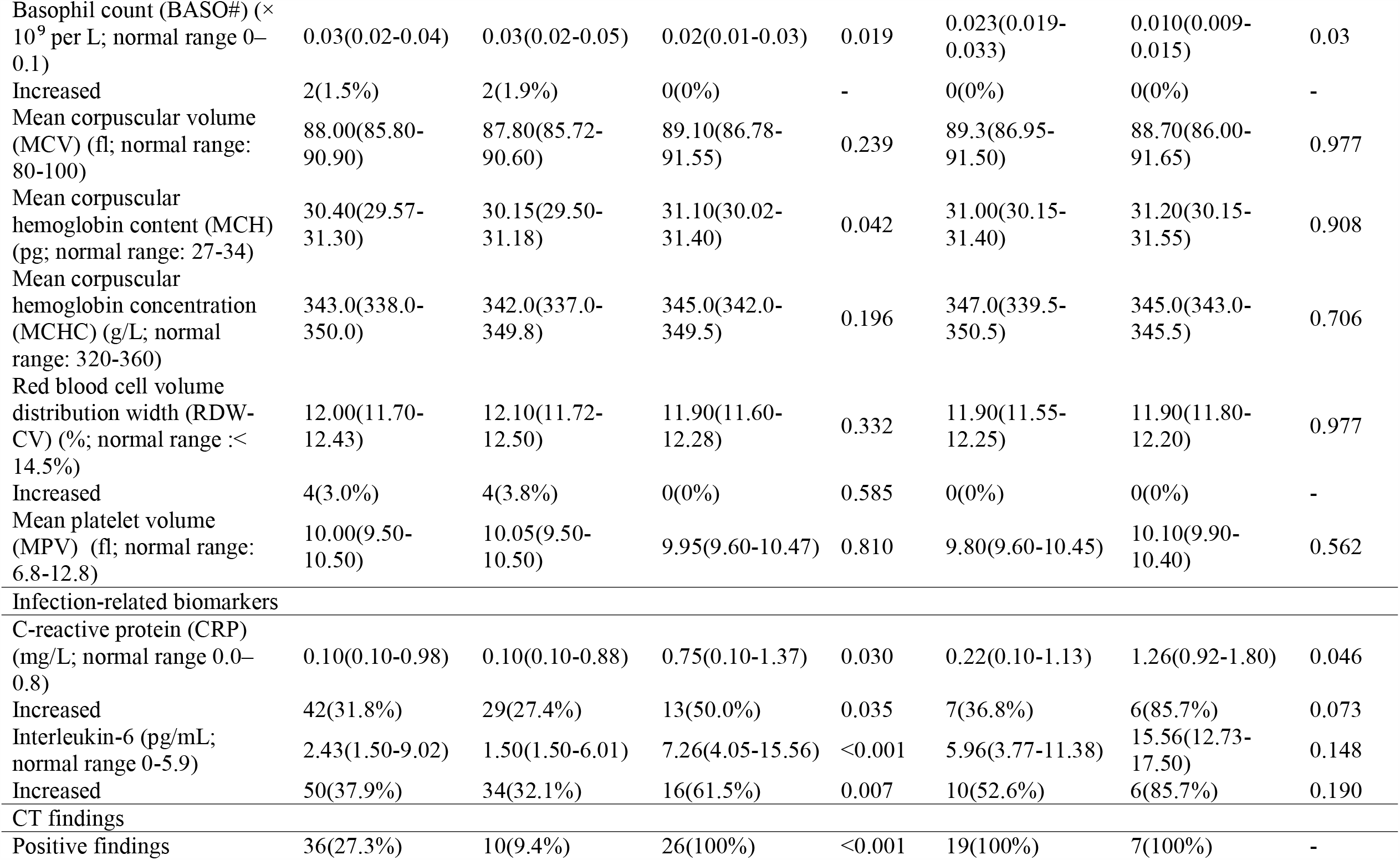
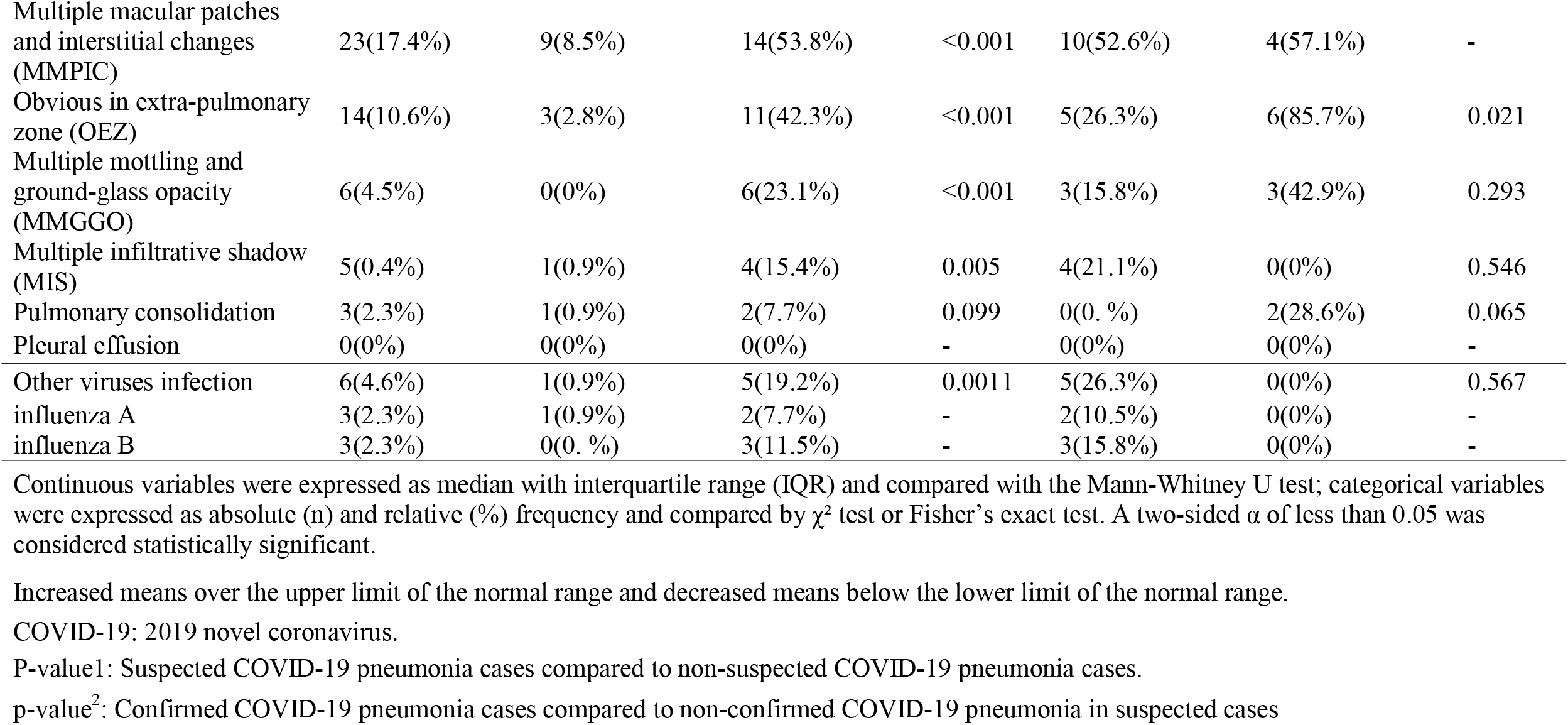
Laboratory results and CT findings of 132 patients admitted to PLA General Hospital (Jan 14–Feb 9, 2020) with the epidemiological history of exposure to COVID-19 in development cohort…

On admission, 26 (100%) and 10 (9.4%) patients had positive CT findings in S-COVID-19-P and N-S-COVID-19-P cases, respectively. In S-COVID-19-P cases, multiple macular patches and interstitial changes accounted for 53.8% (n=14), and multiple mottling and ground-glass opacity accounted for 8.5% (n=9). Positive CT findings in 11 (42.3%) S-COVID-19-P cases and 6 (85.7%) C-COVID-19-P cases were obvious in the extra-pulmonary zone. (Table 3)

The descriptions and statistics of the development cohort’s demographics, baseline, and clinical characteristics were summarized in Table 2, the laboratory results and CT findings were summarized in Table 3. Meanwhile, the same details of the validation cohort, a total of 33 unique admissions with the epidemiological history of exposure to COVID-19 from Feb 10 to Feb 26, 2020, were summarized in Table S2 and Table S3.

### Features selection

Candidate features and univariable associated with S-COVID-19-P were listed in Table S4 from the resulting coefficients of LASSO regularized logistic regression. Therefore, final selected features for model development included: 1) 1 variable of demographic information (age); 2) 4 variables of vital signs [*e*.*g*., Temperature (TEM), Heart rate (HR), etc.]; 3) 5 variables of blood routine values [*e*.*g*., Platelet count (PLT), Monocyte ratio (MONO%), Eosinophil count (EO#), etc.]; 4) 7 variables of clinical signs and symptoms [*e*.*g*., Fever, Fever classification, Shiver, etc.]; 5) 1 infection-related biomarkers [Interleukin-6 (IL-6)]. The final selected features list was shown in Table 4.

**Table 4:** Final selected features for model development

|  |  |  |  |  |
| --- | --- | --- | --- | --- |
| Groups | Final selected features |  |  |  |
| Demographic information | Age |  |  |  |
| Vital signs | Temperature (TEM) | Heart rate (HR) | Diastolic blood pressure (DIAS_BP) | Systolic blood pressure (SYS_BP) |
| Blood routine values | Basophil count (BASO#) | Platelet count (PLT) | Mean corpuscular hemoglobin content (MCH) | Eosinophil count (EO#) |
|  | Monocyte ratio (MONO %) |  |  |  |
| Clinical signs and symptoms on admission | Fever | Shiver | Shortness of breath | Headache |
|  | Fatigue | Sore throat | Fever classification (FC) |  |
| Infection-related biomarkers | Interleukin-6 (IL-6) |  |  |  |

Fever classification : °C , Normal: <= 37.0; Mild fever: 37.1-38.0; Moderate fever: 38.1-39.0; Severe fever: >=39.1.

### Model performance in development and validation cohort

The diagnosis aid model for S-COVID-19-P early identification on admission performed well in both the development and validation cohort according to all evaluation criteria. For the LASSO regularized logistic regression, we introduced the LASSO penalty from C = 0.25 to 7.5 with a step size = 0.25 in scikit-learn package and found C = 7.0 achieved optimal performance to the AUC in the validation set. In the held-out testing set, we found AUC = 0.8409, F-1 score = 0.5714, precision = 0.4000, recall = 1.0000 and specificity = 0.727. In the validation set, we found AUC = 0.9383, F-1 score = 0.6667, precision = 0.5000, recall = 1.0000 and specificity = 0.778. (Table S1)

### Identifying Feature Importance

We analyzed feature importance from the coefficient weights in the LASSO regularized logistic regression model. The list of feature importance rankings of the diagnosis aid model for S-COVID-19-P early identification in the development cohort is shown in Figure 2. Note that the top 5 important features that strongly associated with S-COVID-19-P were Age (0.1115), IL-6 (0.0880), SYS_BP (0.0868), MONO% (0.0679), and Fever classification (0.0569).

**Figure 2.**
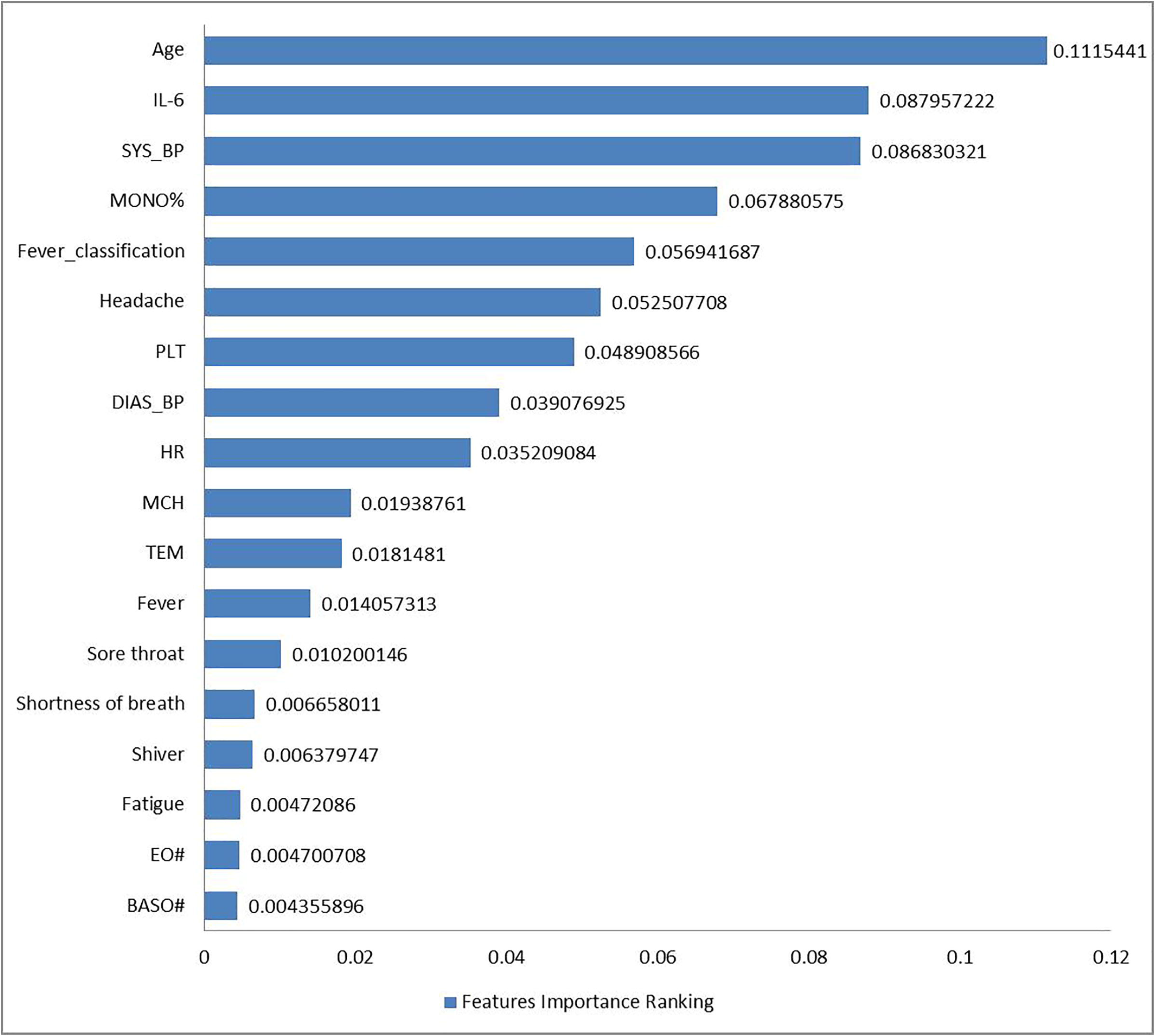
Features Importance Ranking. Feature importance was performed in the development cohort. The associated coefficient weights correspond to the logistic regression model were used for identifying and ranking feature importance. Interleukin-6 (IL-6), Systolic blood pressure (SYS_BP), Monocyte ratio (MONO%), Fever classification (°C, Normal: <= 37.0; mild fever: 37.1-38.0; moderate fever: 38.1-39.0; severe fever: >=39.1), platelet count (PLT), diastolic blood pressure (DIAS_BP), Heart rate (HR), Mean corpuscular hemoglobin content (MCH), Temperature (TEM), Eosinophil count (EO#), Basophil count (BASO#).

### Comparison of diagnostic performance among diagnosis aid model and infection-related biomarkers

The comparison of diagnostic performance among the diagnosis aid model and prominently infection-related biomarkers (lymphopenia, elevated CRP, and elevated IL-6) for early identifying S-COVID-19-P in the development cohort was shown in Table 5. The performance of the diagnosis aid model was better than lymphopenia, elevated CRP, and elevated IL-6, respectively, which resulted in AUCs of 0.841, 0.407, 0.613, and 0.599, Recall of 1.0000, 0.346, 0.500, and 0.615.

**Table 5.** Comparison of diagnostic performance among diagnosis aid model and infection-related biomarkers

|  | Diagnosis aid model | Lymphopenia (<1.0×10<sup>9</sup>/L) | Elevated CRP (>0.8 mg/L) | Elevated IL-6 (>5.9 pg/mL) |
| --- | --- | --- | --- | --- |
| AUC | **0.841** | 0.407 | 0.613 | 0.599 |
| Recall | **1.000** | 0.346 | 0.500 | 0.615 |
| Specificity | 0.727 | **0.840** | 0.726 | 0.679 |
| Precisions | **0.400** | 0.160 | 0.273 | 0.321 |

AUC: the area under the ROC curve
CRP: C-reactive protein

### Online Suspected COVID-19 Pneumonia Diagnosis Aid System

We made the validated diagnosis aid model by LASSO regularized logistic regression algorithm as the “Suspected COVID-19 pneumonia Diagnosis Aid System” which was publicly available through our online portal at https://intensivecare.shinyapps.io/COVID19/.

## Discussion

In this retrospective observation, we evaluated the development and validation of a diagnostic aid model based on the machine-learning algorithms and clinical data without CT images for S-COVID-19-P early identification. The clinical data comes from the demographic information, routinely clinical signs, symptoms, and laboratory tests before the further CT examination. Therefore, in fever clinics under the epidemic outbreak, such a diagnosis aid model might improve triage efficiency, optimize the medical service process, and save medical resources.

From the results in LASSO regularized logistic regression, though some false alarm may exist, the model can identify 100% of the suspected cases in both the held-out testing set and external validation set. By applying this stringent rule to the clinical diagnosis, it is of our great interest to avoid any missed cases. This suggests that our diagnosis aided system can help doctors decide suspected cases in a highly reliable manner.

According to the analysis of features selection and features importance ranking, the univariable from the most demographic information, clinical signs, symptoms, and blood routine values on admission could not show a remarkable association with S-COVID-19-P, which indicated that they may not be informative and increased the difficulty for early identifying S-COVID-19-P with routinely clinical information. Therefore, it is necessary to integrate all the above nonspecific but important features by machine-learning algorithms for secondary analysis and develop cost-effective diagnosis aid models(22, 23).

The infection-related biomarkers, most prominently lymphopenia, elevated CRP, and IL-6 played a key role in identifying clinical infections. For example, lymphopenia has been included as one of three diagnostic criteria for S-COVID-19-P based on 6th-Guidelines-CNHHC(3, 24, 25). In this study, all of these three biomarkers based on the blood routine test on admission could distinguish S-COVID-19-P from the N-S-COVID-19-P well. According to the comparison of diagnostic performance among the diagnosis aid model and these biomarkers, the diagnosis aid model significantly outperformed in AUC and Recall than other biomarkers, which highlighted its potential use for clinical triage. Moreover, we also found that the early elevated monocyte ratio in the development cohort and the early elevated monocyte count could identify S-COVID-19-P from N-S-COVID-19-P well in this study, which suggested that monocyte ratio or monocyte count would also be a new potentially infection-related biomarker for S-COVID-19-P early identification(25).

Although CT scan has become a major diagnostic tool helping for early screening of S-COVID-19-P cases, it could not meet the needs of all patients when the medical resources are insufficient in the epidemic outbreak. From the result of CT findings in the development and validation cohort, there were only 10 (9.4%) and 4 (14.8%) N-S-COVID-19-P cases have mild CT findings on admission, which indicated that the triage strategies for CT scans mainly based on fever or lymphopenia need to be further optimized (26). Therefore, it is meaningful to use machine-learning algorithms to comprehensively analyze clinical symptoms, routine laboratory tests, and other clinical information before further CT examination and develop a diagnosis aid model to improve the triage strategies in fever clinics, which would make a good balance between standard medical principles and limited medical resources.

The developed and validated model performances confirmed that the early identification of S-COVID-19-P in fever clinics could be accurately triaged based only on clinical information without CT images on admission. After feature selection, the final developed model based on fewer predictors could perform well according to most evaluation criteria and also have a better result in further validation. Therefore, the final model based on a small number of features would be likely applicable in most fever clinics.

One of the most effective strategies to control the epidemic outbreak was the establishment of an efficient triaging process for early identification S-COVID-19-P in fever clinics(26). Based on our successful experience in Beijing and well-performed ‘Suspected COVID-19 Pneumonia Diagnosis Aid System’, we have designed the following improved S-COVID-19-P early identification strategies in adult fever clinics (Figure 3). All patients with fever, sore throat, or cough, whether there is hypoxia or not, we proposed routinely take the measurements of blood routine, CRP, IL-6, and influenza virus (A+B) test. Then, if the results of the above tests are normal and the patient without any epidemiological history, home quarantine, regular treatment (such as oral antibiotics), and continuous monitoring clinical signs and symptoms are suggested. If not, a rapid and artificial intelligence assisted evaluation of all clinical results will be required based on our ‘Suspected COVID-19 Pneumonia Diagnosis Aid System’ for S-COVID-19-P early identification, which helps for a decision-support of whether the next CT examination is needed. When the clinical symptoms do not relieve in a few days for home-quarantine patients, they would be required to return for further examination (such as the CT scan). Meanwhile, patients with negative CT findings would also be advised to have a home quarantine with regular treatment and continuous monitoring. Therefore, an artificial intelligence-assisted diagnosis aid system for S-COVID-19-P would take the most advantages of clinical symptoms, routine laboratory tests, and other clinical information available on admission before further CT examination to improve the triage strategies in fever clinics and make a balance between standard medical principles and limited medical resources.

**Figure 3.**
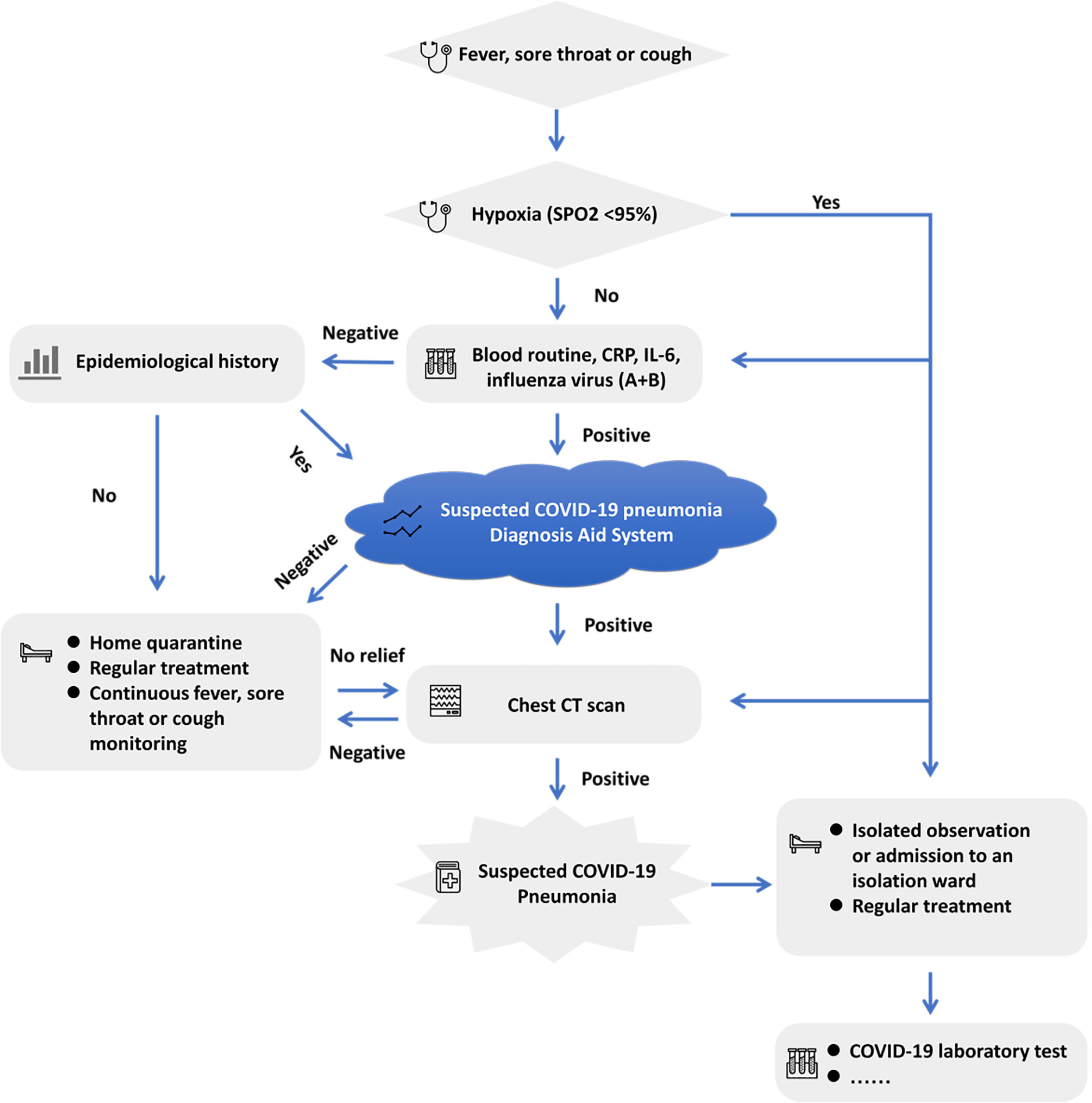
Flow chart for improved S-COVID-19-P early identification strategies in adult fever clinics in PLAGH, China. CRP= C-reactive protein, IL-6= Interleukin-6.

Our current study has several strengths. First, we successfully used a machine-learning algorithm to analyze clinical datasets without CT images and developed a diagnosis aid model for early identification of S-COVID-19-P cases in the fever clinic, which would become a key method to answer the questions of insufficient medical resources in the epidemic outbreak. Second, we integrated most of the routinely available data on admission, including 46 features that are considered to contain the most predictors. Third, we found that the admitted monocyte ratio or monocyte count in blood routine test was more discriminant in S-COVID-19-P cases which might be a new potential infection-related biomarker for early identification. Fourth, we also discussed an optimized triage strategy in fever clinics for early identification of S-COVID-19-P with the help of our new diagnosis aid model which would help to make a balance between standard medical principles and limited medical resources. Fifth, the final model based on a small number of features is likely available in most fever clinics, which has the advantages to increase the possibility of worldwide use and generalizability. Lastly, the developed and validated diagnosis aid model was publicly available as an online triage calculator. This is the first of this method and provides a platform and useful tool for future biomarker and S-COVID-19-P early identification studies in limited-resource settings.

Although the diagnosis results are highly reliable according to the recall score, inevitable limitations may still exist in this study. First, we only evaluated lymphopenia, elevated CRP, and elevated IL-6, while other biomarkers might be more discriminant. Second, the data size was relatively small based on only a single-center fever clinic, which calls for ‘big data’ analysis depend on multiple-center fever clinics. Third, the model was developed and validated in mildly ill patients and with fewer comorbidities; therefore, more well-performing models would be welcomed for a specifical subpopulation. Fourth, since the model was developed and validated in a single-center fever clinic, the performance might vary when evaluated in other fever clinics, particularly if they differ in patient characteristics and COVID-19 prevalence. Therefore, the diagnosis aid model of this study requires further external validation based on different background populations. Fifth, there is a potential risk for misuse of the online calculator. To make the right choice and decision, more consideration should be taken in suited patients and classification threshold (27). Last but not the least, the “Suspected COVID-19 pneumonia Diagnosis Aid System” would only be used as one of the auxiliary references for making clinical and management decisions.

## Conclusion

We successfully used a machine-learning algorithm to develop a diagnosis aid model without CT images for early identification of S-COVID-19-P, and the diagnostic performance was better than lymphopenia, elevated CRP, and elevated IL-6 on admission. The recall score on both held-out testing and validation sets are 100%, suggesting that the model is highly reliable for clinical diagnosis. We also discussed an optimized triage strategy in fever clinics for early identification of S-COVID-19-P with the help of our new diagnosis aid model which would make a good balance between standard medical principles and limited medical resources. To facilitate further validation, the developed diagnosis aid model is available online as a triage calculator.

## Data Availability

The data that support the findings of this study are available from the corresponding author on reasonable request. Participant data without names and identifiers will be made available after approval from the corresponding author, PLAGH and National Health Commission. After publication of study findings, the data will be available for others to request. The research team will provide an email address for communication once the data are approved to be shared with others. The proposal with detailed description of study objectives and statistical analysis plan will be needed for evaluation of the reasonability to request for our data. The corresponding author, PLAGH and National Health Commission will make a decision based on these materials. Additional materials may also be required during the process.

## Author Contributions

I. Conception and design: Cong Feng, Lili Wang, Wei Chen, and Tanshi Li.
II. Administrative support: Juan Zhang, Fei Pan, Li Chen, Zhihong Zhu, Hongju Xiao, Yu Liu, Gang Liu, Wei Chen, and Tanshi Li.
III. Provision of study materials or patients: Hua Chen, Yingchan Wang, Xiangzheng Su,Sai Huang
IV. Collection and assembly of data: Lin Tian, Weixiu Zhu, Wenzheng Sun, Liping Zhang, Qingru Han.
V. Data analysis and interpretation: Cong Feng, Lili Wang, Wei Chen, and Tanshi Li.
VI. Manuscript writing: All authors
VII. Final approval of manuscript: All authors

## Conflicts of Interest

The authors declare that they have no conflict of interest.

## Data sharing

The data that support the findings of this study are available from the corresponding author on reasonable request. Participant data without names and identifiers will be made available after approval from the corresponding author, PLAGH, and National Health Commission. After the publication of the study findings, the data will be available for others to request. The research team will provide an email address for communication once the data are approved to be shared with others. The proposal with a detailed description of study objectives and a statistical analysis plan will be needed for evaluation of the reasonability to request for our data. The corresponding author, PLAGH, and National Health Commission will make a decision based on these materials. Additional materials may also be required during the process.

## Reporting Checklist

The authors have completed the STROBE reporting checklist.

---

**Table S1:** Comparison of different algorisms

| Algorithms/Performance | Cohorts | AUC | F-1 score | Precisions | Recall | Specificity |
| --- | --- | --- | --- | --- | --- | --- |
| Logistic regression with LASSO | Development cohort | **0.841** | **0.571** | **0.400** | **1.000** | 0.727 |
|  | Validation cohort | **0.938** | **0.667** | **0.500** | **1.000** | 0.778 |
| logistic regression with Ridge regularization | Development cohort | 0.796 | 0.462 | 0.333 | 0.750 | 0.727 |
|  | Validation cohort | 0.864 | 0.571 | 0.400 | **1.000** | 0.667 |
| Decision tree | Development cohort | 0.580 | 0.286 | 0.333 | 0.250 | 0.909 |
|  | Validation cohort | 0.500 | 0.000 | 0.000 | 0.000 | **1.000** |
| Adaboost algorithms | Development cohort | 0.500 | 0.000 | 0.000 | 0.000 | **0.818** |
|  | Validation cohort | 0.790 | 0.222 | 0.333 | 0.167 | 0.926 |

AUC: the area under the ROC curve

**Table S2:**
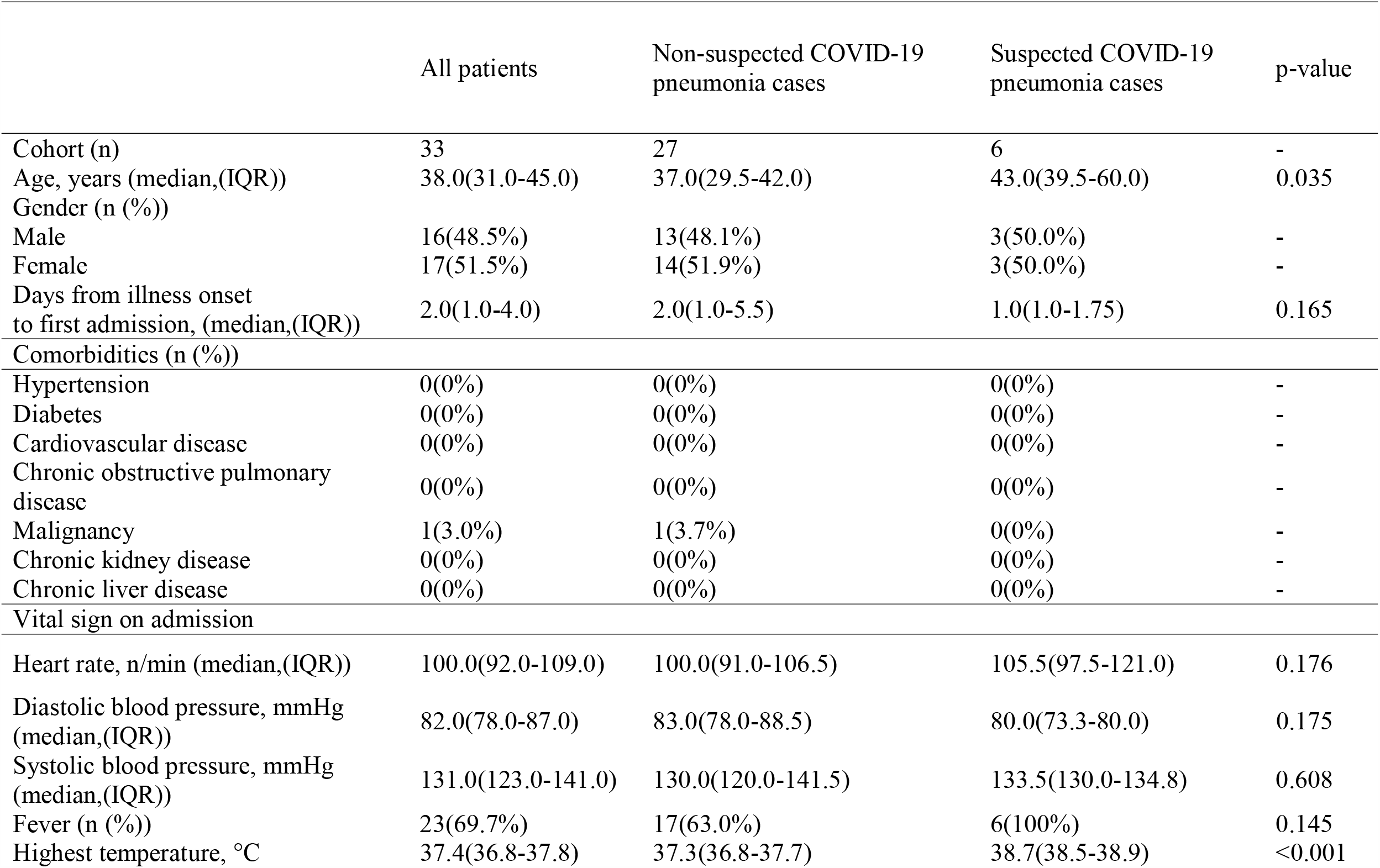
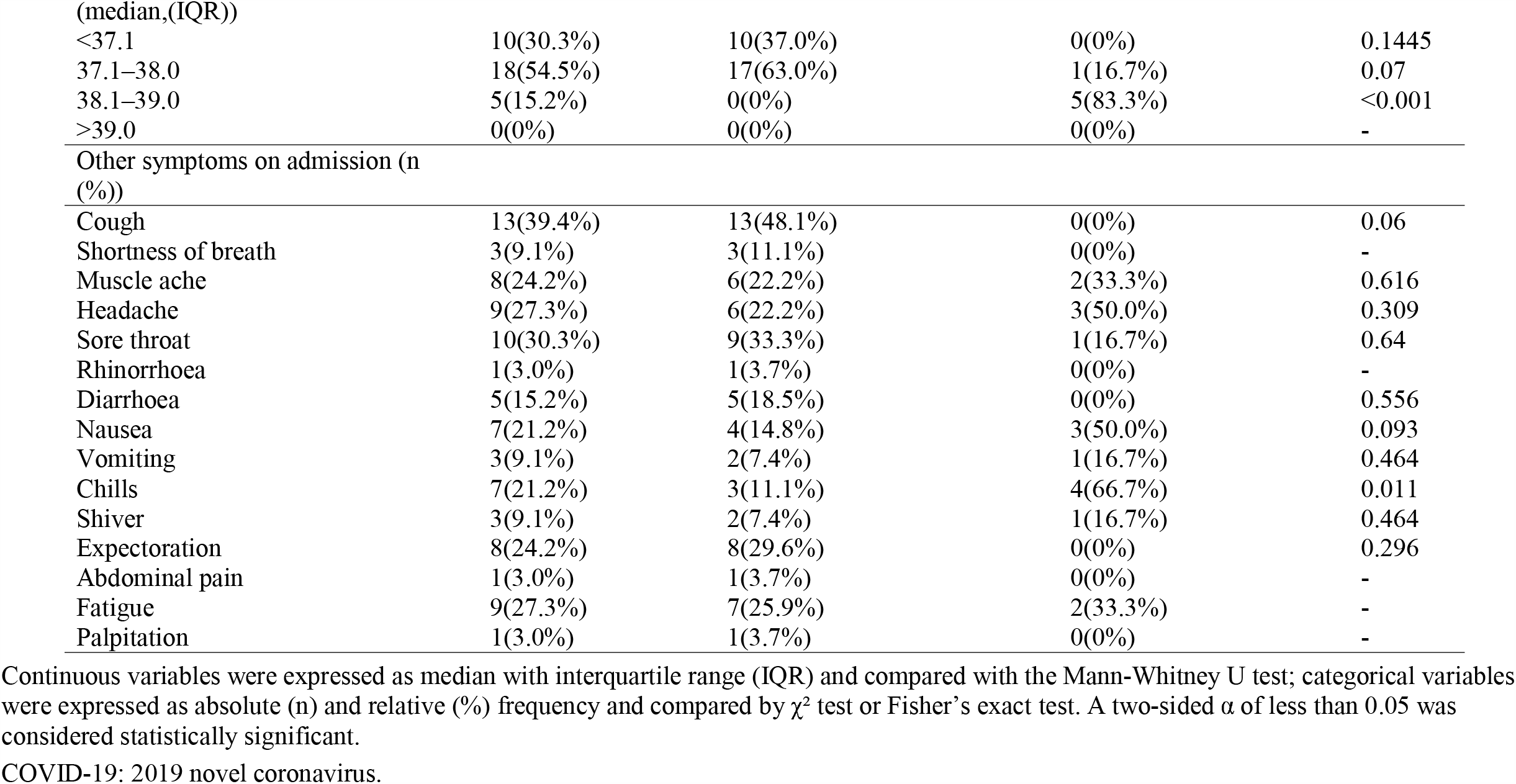
Demographics, baseline and clinical characteristics of 33 patients admitted to PLA General Hospital (Feb 10–Feb 26, 2020) with the epidemiological history of exposure to COVID-19 in validation cohort.

**Table S3:** Laboratory results and CT findings of 33 patients admitted to PLA General Hospital (Feb 10–Feb 26, 2020) with the epidemiological history of exposure to COVID-19 in validation cohort…

|  | All patients | Non-suspected<br>COVID-19<br>pneumonia cases | Suspected COVID-19<br>pneumonia cases | p-value |
| --- | --- | --- | --- | --- |
| Cohort (n) | 33 | 27 | 6 | - |
| Blood routine values |  |  |  |  |
| White blood cell count (WBC) ($\times 10^9$ per L; normal range 3.5–10.0) | 6.78(5.36-8.62) | 6.56(5.31-7.79) | 8.89(7.95-9.82) | 0.025 |
| Increased | 3(9.1%) | 1(3.7%) | 2(33.3%) | 0.078 |
| Decreased | 0(0%) | 0(0%) | 0(0%) | - |
| Red blood cell count (RBC) ($\times 10^{12}$ per L; normal range: male 4.3–5.9, female 3.9–5.2) | 4.64(4.16-5.05) | 4.74(4.33-5.20) | 4.34(4.12-4.61) | 0.08 |
| Decreased | 2(6.1%) | 1(3.7%) | 1(16.7%) | 0.335 |
| Hemoglobin (HGB) (g/L; normal range: male 137.0–179.0, female 116.0–155.0) | 142.0(130.0-151.0) | 143.0(133.0-152.5) | 131.5(128.0-138.0) | 0.088 |
| Decreased | 2(6.1%) | 1(3.7%) | 1(16.7%) | 0.335 |
| Hematocrit (HCT) (normal range: male 0.4–0.52, female 0.37–0.47) | 0.41(0.37-0.44) | 0.42(0.38-0.45) | 0.37(0.37-0.38) | 0.059 |
| Increased | 1(3.0%) | 1(3.7%) | 0(0%) | - |
| Decreased | 13(39.4%) | 8(29.6%) | 5(83.3%) | 0.025 |
| Platelet count (PLT) ($\times 10^9$ per L; normal range 100.0–300.0) | 231.0(200.0-261.0) | 231.0(201.5-276.5) | 234.0(206.8-242.5) | 0.834 |
| Decreased | 1(3.0%) | 1(3.7%) | 0(0%) | - |
| Lymphocyte ratio (LYMPH %) (0.2-0.4) | 0.19(0.14-0.29) | 0.22(0.17-0.30) | 0.11(0.09-0.13) | 0.001 |
| Increased | 1(3.0%) | 1(3.7%) | 0(0%) | - |
| Decreased | 18(54.5%) | 12(44.4%) | 6(100.0%) | 0.021 |
| Lymphocyte count (LYMPH#) ($\times 10^9$ per L; normal range 1.0–4.0) | 1.36(1.01-1.87) | 1.46(1.21-1.96) | 1.00(0.98-1.01) | 0.005 |
| Increased | 0(0%) | 0(0%) | 0(0%) | - |
| Decreased | 7(21.2%) | 4(14.8%) | 3(50.0%) | 0.093 |
| Neutrophil ratio (NEUT %) (0.5-0.7) | 0.73(0.59-0.78) | 0.71(0.58-0.76) | 0.78(0.75-0.85) | 0.057 |
| Increased | 20(60.6%) | 15(55.6%) | 5(83.8%) | 0.364 |
| Decreased | 3(9.1%) | 3(11.1%) | 0(0%) | - |
| Neutrophil count (NEUT#) ($\times 10^9$ per L; normal range 2.0–7.0) | 4.76(3.07-7.01) | 4.20(3.02-5.78) | 7.29(6.07-8.15) | 0.031 |
| Increased | 9(27.3%) | 5(18.5%) | 4(66.7%) | 0.034 |
| Decreased | 0(0%) | 0(0%) | 0(0%) | - |
| Eosinophil ratio (EO %) (0.01-0.05) | 0.008(0.003-0.025) | 0.008(0.004-0.028) | 0.001(0.0003-0.014) | 0.129 |
| Increased | 4(12.1%) | 3(11.1%) | 1(16.7%) | - |
| Eosinophil count (EO#) ($\times 10^9$ per L; normal range 0.05–0.3) | 0.05(0.02-0.16) | 0.05(0.03-0.16) | 0.01(0.003-0.11) | 0.146 |
| Increased | 4(12.1%) | 3(11.1%) | 1(16.7%) | - |
| Monocyte ratio (MONO %) (0.03-0.08) | 0.06(0.04-0.08) | 0.06(0.04-0.07) | 0.07(0.06-0.10) | 0.154 |
| Increased | 9(27.3%) | 6(22.2%) | 3(50.0%) | 0.309 |
| Monocyte count (MONO#) ($\times 10^9$ per L; normal range 0.12–0.8) | 0.38(0.31-0.46) | 0.36(0.29-0.44) | 0.61(0.55-0.77) | <0.001 |
| Increased | 2(6.1%) | 0(0%) | 2(33.3%) | 0.028 |
| Basophil ratio (BASO %) (0-0.01) | 0.003(0.002-0.006) | 0.003(0.002-0.007) | 0.003(0.001-0.004) | 0.422 |
| Increased | 2(6.1%) | 2(7.4%) | 0(0%) | - |
| Basophil count (BASO#) ($\times 10^9$ per L; normal range 0–0.1) | 0.02(0.01-0.04) | 0.02(0.01-0.04) | 0.02(0.01-0.04) | 0.91 |
| Increased | 0(0%) | 0(0%) | 0(0%) | - |
| Mean corpuscular volume (MCV) (fl; normal range: 80-100) | 87.10(85.60-89.40) | 87.10(85.20-89.65) | 87.45(85.80-88.72) | 0.944 |
| Mean corpuscular hemoglobin content (MCH) (pg; normal range: 27-34) | 30.50(29.50-31.10) | 29.90(29.45-31.05) | 30.80(30.52-31.90) | 0.315 |
| Mean corpuscular hemoglobin concentration (MCHC) (g/L; normal range: 320-360) | 348.0(340.0-354.0) | 347.0(338.0-353.0) | 353.5(347.0-360.0) | 0.215 |
| Red blood cell volume distribution width (RDW-CV) (%; normal range :< 14.5%) | 12.00(11.80-12.70) | 12.00(11.80-12.60) | 12.25(11.60-14.03) | 0.623 |
| Increased | 2(6.1%) | 0(0%) | 2(33.3%) | 0.028 |
| Mean platelet volume (MPV) (fl; normal range: 6.8-12.8) | 9.90(9.60-10.90) | 9.90(9.60-10.90) | 10.10(9.68-10.75) | 0.743 |
| Infection-related biomarkers |  |  |  |  |
| C-reactive protein (CRP) (mg/L; normal range 0.0–5.0) | 0.10(0.10-0.95) | 0.10(0.10-0.19) | 7.56(2.55-8.41) | <0.001 |
| Increased | 9(27.3%) | 4(14.8%) | 5(83.3%) | 0.003 |
| Interleukin-6 (pg/mL; normal range 0.5-9) | 1.50(1.50-20.54) | 1.50(1.50-1.59) | 26.79(21.94-79.94) | <0.001 |
| Increased | 10(30.3%) | 4(14.8%) | 6(100.0%) | <0.001 |
| CT findings |  |  |  |  |
| Positive findings | 10(30.3%) | 4(14.8%) | 6(100%) | <0.001 |
| Multiple macular patches and interstitial changes (MMPIC) | 6(18.2%) | 4(14.8%) | 2(33.3%) | 0.295 |
| Obvious in extra-pulmonary zone (OEZ) | 0(0%) | 0(0%) | 0(0%) | - |
| Multiple mottling and ground-glass opacity (MMGGO) | 1(3.0%) | 0(0%) | 1(16.7%) | 0.182 |
| Multiple infiltrative shadow (MIS) | 4(12.1%) | 0(0%) | 4(100.0%) | <0.001 |
| Pulmonary consolidation | 3(9.1%) | 0(0%) | 3(50.0%) | 0.004 |
| Pleural effusion | 1(3.0%) | 0(0%) | 1(16.7%) | 0.182 |
| Other viruses infection | 0(0. %) | 0(0. %) | 0(0. %) | - |
| influenza A | 0(0. %) | 0(0. %) | 0(0. %) | - |
| influenza B | 0(0. %) | 0(0. %) | 0(0. %) | - |

Continuous variables were expressed as median with interquartile range (IQR) and compared with the Mann-Whitney U test; categorical variables were expressed as absolute (n) and relative (%) frequency and compared by $\chi^2$ test or Fisher's exact test. A two-sided $\alpha$ of less than 0.05 was considered statistically significant.
Increased means over the upper limit of the normal range and decreased means below the lower limit of the normal range.
COVID-19: 2019 novel coronavirus.

**Table S4:** Candidate features and univariable association with S-COVID-19-P

| Candidate features | Association and weight |
| --- | --- |
| Age | 0.1115441 |
| IL-6 | 0.087957222 |
| SYS_BP | 0.086830321 |
| MONO% | 0.067880575 |
| Fever_class | 0.056941687 |
| Headache | 0.052507708 |
| DIAS_BP | 0.039076925 |
| HR | 0.035209084 |
| MCH | 0.01938761 |
| TEM | 0.0181481 |
| Fever | 0.014057313 |
| Sore throat | 0.010200146 |
| WBC | 0 |
| LYMPH% | 0 |
| LYMPH# | 0 |
| Chills | 0 |
| MONO# | 0 |
| EO% | 0 |
| BASO% | 0 |
| NEUT% | 0 |
| HCT | 0 |
| MCV | 0 |
| MCHC | 0 |
| RDW-CV | 0 |
| MPV | 0 |
| CRP | 0 |
| NEUT# | 0 |
| DOA | 0 |
| Rhinorrhoea | 0 |
| Muscle ache | 0 |
| HGB | 0 |
| Gender | 0 |
| Diarrhoea | 0 |
| Cough | 0 |
| Palpitation | 0 |
| RBC | 0 |
| Abdominal pain | 0 |
| Vomiting | 0 |
| Nausea | 0 |
| Expectoration | 0 |
| BASO# | -0.004355896 |
| EO# | -0.004700708 |
| Fatigue | -0.00472086 |
| Shiver | -0.006379747 |
| Shortness of breath | -0.006658011 |
| PLT | -0.048908566 |

The abbreviations were referenced from the Table 1.

## Notes

### Competing Interest Statement

The authors have declared no competing interest.

### Summary of Updates

We updated the author list, author contribution, and funding information.

## References

1. Wu F, Zhao S, Yu B et al. A new coronavirus associated with human respiratory disease in China. Nature 2020.

2. Huang C, Wang Y, Li X et al. Clinical features of patients infected with 2019 novel coronavirus in Wuhan, China. Lancet 2020; 395: 497–506.

3. Chen N, Zhou M, Dong X et al. Epidemiological and clinical characteristics of 99 cases of 2019 novel coronavirus pneumonia in Wuhan, China: a descriptive study. Lancet 2020; 395: 507–513.

4. Chan JF, Yuan S, Kok KH et al. A familial cluster of pneumonia associated with the 2019 novel coronavirus indicating person-to-person transmission: a study of a family cluster. Lancet 2020; 395: 514–523.

5. Xu Z, Shi L, Wang Y et al. Pathological findings of COVID-19 associated with acute respiratory distress syndrome. Lancet Respir Med 2020.

6. Kim JY, Choe PG. The First Case of 2019 Novel Coronavirus Pneumonia Imported into Korea from Wuhan, China: Implication for Infection Prevention and Control Measures. 2020; 35: e61.

7. Wang C, Horby PW, Hayden FG et al. A novel coronavirus outbreak of global health concern. Lancet 2020; 395: 470–473.

8. The L. Emerging understandings of 2019-nCoV. Lancet 2020; 395: 311.

9. Chang, Lin M, Wei L et al. Epidemiologic and Clinical Characteristics of Novel Coronavirus Infections Involving 13 Patients Outside Wuhan, China. Jama 2020.

10. Holshue ML, DeBolt C, Lindquist S et al. First Case of 2019 Novel Coronavirus in the United States. N Engl J Med 2020; 382: 929–936.

11. Lee EYP, Ng MY, Khong PL. COVID-19 pneumonia: what has CT taught us? Lancet Infect Dis 2020.

12. Shi H, Han X, Jiang N et al. Radiological findings from 81 patients with COVID-19 pneumonia in Wuhan, China: a descriptive study. Lancet Infect Dis 2020.

13. Rajkomar A, Dean J, Kohane I. Machine Learning in Medicine. N Engl J Med 2019; 380: 1347–1358.

14. Bailly S, Meyfroidt G, Timsit JF. What’s new in ICU in 2050: big data and machine learning. 2018; 44: 1524–1527.

15. Raita Y, Goto T, Faridi MK et al. Emergency department triage prediction of clinical outcomes using machine learning models. Crit Care 2019; 23: 64.

16. Tomar A, Gupta N. Prediction for the spread of COVID-19 in India and effectiveness of preventive measures. Sci Total Environ 2020; 728: 138762.

17. Chimmula VKR, Zhang L. Time Series Forecasting of COVID-19 transmission in Canada Using LSTM Networks. Chaos Solitons Fractals 2020; 135: 109864.

18. Ayyoubzadeh SM, Ayyoubzadeh SM. Predicting COVID-19 Incidence Through Analysis of Google Trends Data in Iran: Data Mining and Deep Learning Pilot Study. 2020; 6: e18828.

19. Reid S, Tibshirani R. Regularization Paths for Conditional Logistic Regression: The clogitL1 Package. J Stat Softw 2014; 58.

20. Bradley APJPr. The use of the area under the ROC curve in the evaluation of machine learning algorithms. 1997; 30: 1145–1159.

21. Steyerberg EW, Vickers AJ, Cook NR et al. Assessing the performance of prediction models: a framework for traditional and novel measures. Epidemiology 2010; 21: 128–138.

22. Henry KE, Hager DN, Pronovost PJ et al. A targeted real-time early warning score(TREWScore) for septic shock. Sci Transl Med 2015; 7: 299ra122.

23. Komorowski M, Celi LA. The Artificial Intelligence Clinician learns optimal treatment strategies for sepsis in intensive care. 2018; 24: 1716–1720.

24. Wong CK, Lam CW, Wu AK et al. Plasma inflammatory cytokines and chemokines in severe acute respiratory syndrome. Clin Exp Immunol 2004; 136: 95–103.

25. Wu J, Wu X, Zeng W et al. Chest CT Findings in Patients with Corona Virus Disease 2019 and its Relationship with Clinical Features. Invest Radiol 2020.

26. Zhang J, Zhou L, Yang Y et al. Therapeutic and triage strategies for 2019 novel coronavirus disease in fever clinics. Lancet Respir Med 2020.

27. Flechet M, Guiza F, Schetz M et al. AKIpredictor, an online prognostic calculator for acute kidney injury in adult critically ill patients: development, validation and comparison to serum neutrophil gelatinase-associated lipocalin. Intensive Care Med 2017; 43: 764–773.

